# *“Taking PrEP every day for me is a challenge”:* Barriers, and facilitators to accessing HIV pre-exposure prophylaxis (PrEP) services among young people in Gauteng, South Africa

**DOI:** 10.64898/2026.01.15.26344177

**Authors:** C. Mongwenyana-Makhutle, C. Hendrickson, S. Dubazana, J. M. Mazibuko, R. Motaung, N. Mokhesi, S Bokolo, C Chetty-Makkan, A. Moolla, L. Long, J. Miot

## Abstract

**Background:** Pre-exposure prophylaxis (PrEP) is an effective HIV prevention strategy, yet uptake remains suboptimal and ensuring access to PrEP services among young people is critical. We explored the barriers and facilitators associated with accessing PrEP among young people

**Methods:** Focus group discussions (FGDs) were conducted in April and May 2023 in the City of Johannesburg district in South Africa. Participants were adult males and females (ages 18-35) self-reporting as HIV negative with or without previous PrEP use and exposure. We analysed transcripts using a deductive and inductive thematic approach. Two transcripts were coded by three coders to test reliability and saturation was reached when no new themes emerged.

**Results:** Findings were mapped to the Socio-Ecological Model (SEM). Key barriers emerged across levels. Individual level barriers included difficulty adhering to daily oral PrEP and fear of needles. Interpersonal challenges included anxiety about disclosing PrEP use to partners or family. Institutional and organisational barriers involved long clinic queues, negative staff attitudes, limited confidentiality and inadequate PrEP information. At the community level, stigma and misconceptions linking PrEP to HIV treatment deterred uptake. Several facilitators emerged. Individual motivation to remain HIV negative was a strong driver of uptake. Supportive relationships and open communication with friends and family enhanced acceptance. Organisational enablers included access to clear information, youth-friendly services, competent providers and delivery options such as home delivery or pharmacy access. Community awareness initiatives reduced stigma and structural support in a form of free PrEP improved affordability and access.

**Conclusion:** Young people’s access to PrEP is shaped by interactions across individual, relational, institutional, community, and structural levels. Tailored interventions that address personal barriers, strengthen supportive environments, improve service delivery, and ensure affordability are likely to strengthen uptake and adherence. These findings highlight the need for multilevel strategies to optimize PrEP implementation among youth.

## Background

Globally in 2023, approximately 39 million people were living with the Human Immunodeficiency Virus (HIV), over 20% of whom (8.3 million) lived in South Africa[1]. Despite significant progress in HIV prevention programming over the past several years, South Africa still accounts for one-fifth (20%) of new HIV infections globally[2, 3]. To address this, the World Health Organization (WHO) recently endorsed the 2022–2030 Global Health Sector Strategy (GHSS) under the United Nations Sustainable Development Goal (UN-SDGs-3), which aims to target HIV control and prevention by reducing the global HIV burden by at least 50% in high-burdened countries by 2030[4].

Pre-exposure prophylaxis (PrEP), approved by the WHO for use among all people vulnerable to HIV infection, is an effective biomedical intervention for HIV prevention and holds significant promise for reducing new HIV infections[5, 6]. When taken daily, PrEP significantly decreases the risk of contracting HIV, with an effectiveness rate of over 99% if taken consistently and as prescribed[7]. South Africa adopted the WHO’s recommendation to offer daily oral PrEP to those at “substantial risk of HIV infection” in 2016, beginning the PrEP rollout to select vulnerable populations at pilot sites and expanding nationally from 2019[5]. South Africa also has several ongoing demonstration projects that are assessing the feasibility of the national introduction of the dapivirine ring and the cabotegravir long-acting injectable, two additional formulations of PrEP[8].

Daily oral PrEP is a self-administered modality and holds particular promise as an HIV prevention method for young people, particularly among young women who may have limited ability to negotiate condom use[9]. However, challenges with uptake, initiation and adherence remain widespread despite oral PrEP being readily available in public health clinics[10]. Since its rollout in 2016, oral PrEP provision has been scaled up and, as of December 2024, almost 1.8 million individuals were initiated on oral PrEP across 4,291 facilities in South Africa[11]. Despite substantial national scale-up, the uptake and sustained use of oral PrEP among young people has not been optimal for disease control[10]. Young people cite accessibility and availability as challenges to uptake, as well as poor interactions with health care providers, particularly judgmental attitudes and lack of confidentiality[12–14]. In addition, young people are also reluctant to initiate PrEP due to low HIV risk perception, concerns about side effects and stigma, and lack of partner or parental support[14–16].

In addition to the individual- and community-level barriers to oral PrEP uptake and persistence described above, there is increasing concern about the sustainability of HIV prevention programmes in the context of declining international funding[17]. The recent withdrawal of funding has resulted in the closure of clinics and community services focused on the most vulnerable populations in South Africa. It is estimated that the withdrawal of PEPFAR funding could increase to expected infections of between 29% and 57% and AIDS-related deaths by between 33% and 38%, respectively[18]. Not only do these projections highlight the precarious state of HIV prevention infrastructure, they also indicate how funding cuts threaten access to HIV prevention services critical to youth and young women.

In South Africa, where the HIV epidemic remains a significant public health concern, ensuring access to PrEP services among young people is critical. We conducted a study in Gauteng to explore the barriers and facilitators associated with accessing PrEP among young people with diverse exposure to PrEP (i.e., PrEP naïve and experienced) to gain insights on factors that enhance or hinder their ability to effectively utilize this preventive strategy.

## Methods

### Study design and setting

We enrolled participants in Johannesburg, Gauteng, between April 2023 and May 2023. The data was collected from either a community council office or one of three community health centres within the City of Johannesburg district in Gauteng, South Africa. We conducted focus group discussions (FGDs) to facilitate conversations around accessing PrEP. We stratified these FGDs by gender (male or female) and prior use of daily oral PrEP (PrEP experienced versus PrEP naïve). In this study, we define PrEP-naïve as someone who has never taken PrEP and PrEP-experienced as someone who is using or has used PrEP before^31^.

### Study population and sampling

We used a purposive sampling approach to recruit potential participants from Indlela’ s Behavioural Hub (B-Hub), a research cohort developed by Indlela, a behavioural economics unit within the Health Economics and Epidemiology Research Office (HE²RO)[14]. At the time of study recruitment, the B-Hub consisted of 500 members from various local communities around the City of Johannesburg (COJ) in Gauteng province (South Africa) who had consented and enrolled for behavioural studies. Upon enrolment into the B-Hub, participants completed a demographic and health survey which enabled the research team to develop a screening sampling frame for our study. From this frame, we purposively identified potential participants who met the study criteria. Eligible individuals included males and females aged 18-35 years who self-reported to be HIV negative and who were either PrEP naïve or experienced, to capture a diverse range of perceptions and experiences with PrEP use. Identified participants were contacted telephonically by a study member and invited to participate in an FGD. During this call, participants interested in taking part in the study FGD were screened for eligibility. Those who met the criteria were invited to attend an FGD session. On the day of the FGD, all potential participants were re-screened in person to confirm eligibility.

### Data collection procedures

We conducted in-person FGDs in private and secure spaces; discussions were facilitated by a trained, experienced researcher fluent in English and other South African local languages including Sesotho, Setswana and IsiZulu. All participants provided written informed consent and audio recording consent on the day of the FGD. We collected some basic sociodemographic information from each participant before beginning the FGD; these questions comprised sex assigned at birth, current age, preferred language, education, employment and prior exposure to PrEP.

In addition to the primary researcher, two researchers were present during the FGDs with both assuming the role of notetaker and observer and, in some instances, co-facilitator. We used a semi-structured interview guide with suggested probes to encourage participation in the FGDs (Appendix A). The guide was designed by the research team, drawing on insights from similar studies. It explored basic PrEP knowledge and then focused on preferences around the characteristics of PrEP service delivery including cost, cadre of healthcare providers, refill frequency, integration of services, PrEP modality as well as barriers and facilitators to accessing PrEP at various service delivery points. Educational material with information on what PrEP does, the different PrEP modalities that are available or currently in development, service points to access PrEP and online resources where further information on PrEP could be obtained were shown to participants to assist in facilitating the discussion with PrEP naïve individuals during the session. Educational material was provided to participants after assessing their PrEP knowledge and before discussing the barriers and facilitators to accessing PrEP. As part of maintaining anonymity, pseudonyms were assigned to participants and participants were provided with a study identification number captured on the screening form. The FGD duration ranged from 45 to 60 minutes and were all audio-recorded. Participants were reimbursed with ZAR150 ($8.59; 2023 average exchange rate) for their time. The average exchange rate in 2023 was about USD 0.0543 per ZAR 1.

### Data Analysis

We transcribed the FGD recordings verbatim and translated them into English. Three study team members (CM, SD, MM) independently read through the transcripts and applied line-by-line codes to generate the initial codes. To ensure consistency, team members met to discuss their coding and reached consensus on the final set of codes. These consensus-based codes were then categorised into potential themes and sub-themes through iterative discussions, forming a final thematic framework. A codebook was generated from this framework and reviewed by the first author (CM) to ensure that the coding structure and themes accurately reflected the FGD transcripts. No new themes emerged from the data, indicating that saturation had been reached. Transcripts were subsequently imported to NVivo 11 software[19, 20] and then the team assessed inter-coder reliability by independently coding two transcripts using the final codebook, achieving 98-100% agreement score. The data was analysed using both inductive and deductive approaches, with themes interpreted and structured using the Social Ecological Model (SEM)[21, 22]. This model highlights the multi-level interplay of factors influencing PrEP access and use, including individual, interpersonal, institutional, community, organisational and structural factors[21–23].

### Ethical approval

We obtained written informed consent as well as audio recording consent for all study participation. The study was conducted in compliance with regulatory requirements and ethical guidelines for research with human subjects in South Africa and institutional policies (ICH Good Clinical Practice). Ethical oversight was provided by the Human Research Ethics Committee (Medical) at the University of the Witwatersrand (211122), South African National Clinical Trials Registry, registration number: DOH-27-032022-5627 and the Gauteng Department of Health (GP_202201_028).

## Results

We conducted four FGDs with PrEP naïve males (2 FGDs) and females (2 FGDs), and two FGDs with PrEP experienced males (1 FGD) and females (1 FGD), 6 FGDs in total. Participants gave the same responses during the telephonic screening and the re-screening on the day the FGD was conducted. In total, 72 participants were contacted, 32 of whom decided to enrol and participate in our study, while 38 never arrived due to being unreachable or unavailable on the day of the FGDs and 2 were ineligible to participate.

**Figure 1.**
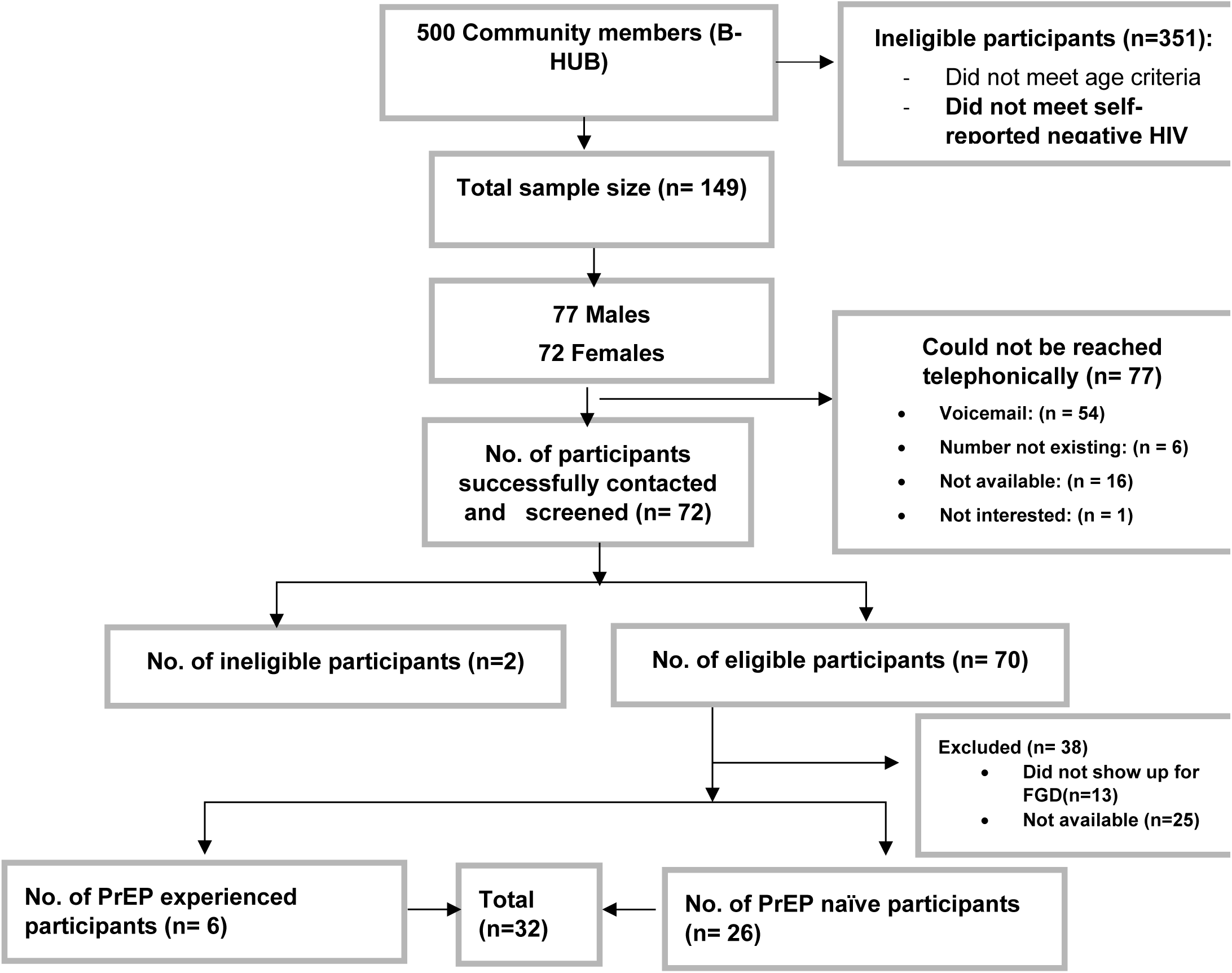
Recruitment consort diagram. The FGDs varied in size, with the smallest being a male PrEP experienced group (n=2) and the largest a male PrEP naïve group (n=11) (Appendix B). Most participants were men (19; 59.3%), unemployed (26; 81.3%), young (median age: 28, IQR: 23–32) and PrEP naïve (26; 81.2%).

**Table 1:**
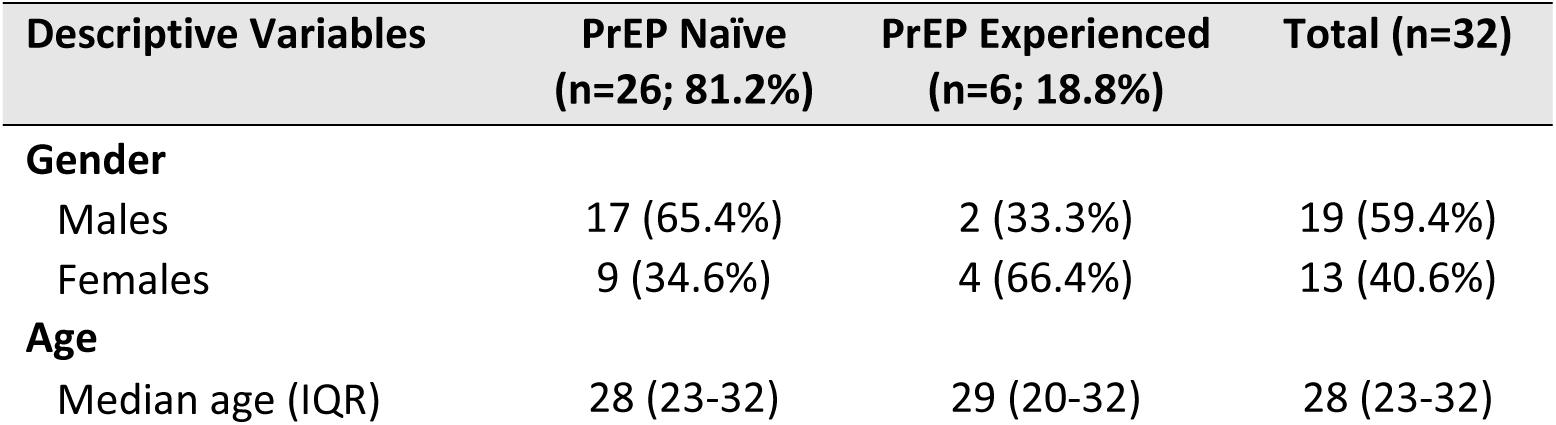
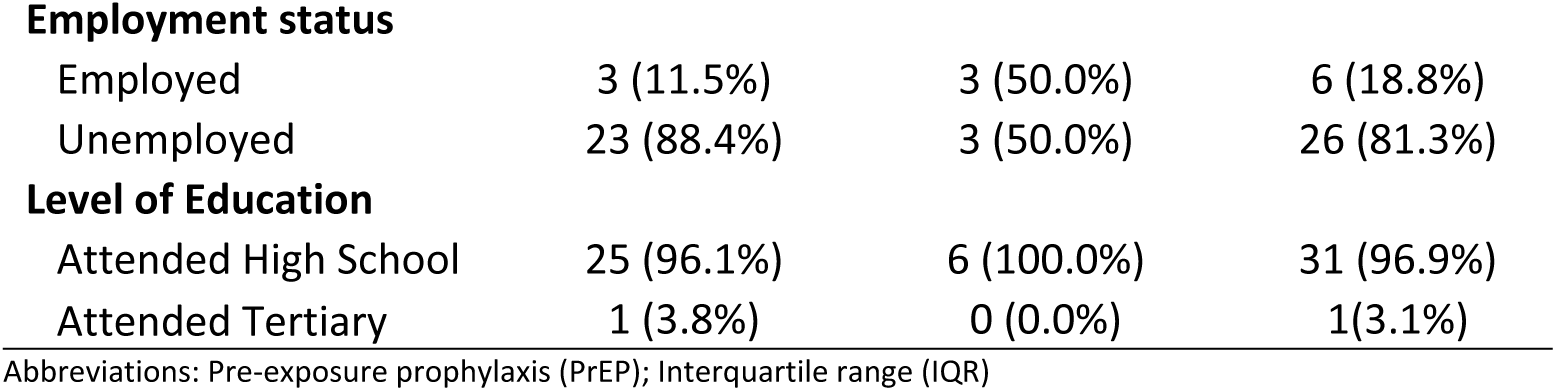
Sample demographics (n=32)

| Descriptive Variables | PrEP Naïve (n=26; 81.2%) | PrEP Experienced (n=6; 18.8%) | Total (n=32) |
| --- | --- | --- | --- |
| **Gender** |  |  |  |
| Males | 17 (65.4%) | 2 (33.3%) | 19 (59.4%) |
| Females | 9 (34.6%) | 4 (66.4%) | 13 (40.6%) |
| **Age** |  |  |  |
| Median age (IQR) | 28 (23-32) | 29 (20-32) | 28 (23-32) |
| **Employment status** |  |  |  |
| Employed | 3 (11.5%) | 3 (50.0%) | 6 (18.8%) |
| Unemployed | 23 (88.4%) | 3 (50.0%) | 26 (81.3%) |
| **Level of Education** |  |  |  |
| Attended High School | 25 (96.1%) | 6 (100.0%) | 31 (96.9%) |
| Attended Tertiary | 1 (3.8%) | 0 (0.0%) | 1 (3.1%) |

Abbreviations: Pre-exposure prophylaxis (PrEP); Interquartile range (IQR)

Our results present the perceived barriers and facilitators of accessing PrEP among young adults across a modified socio-ecological model (Fig 2). This includes individual, interpersonal, institutional, community, organizational, and structural factors. At the individual level, some participants feared side effects, needles or had a concern of forgetting to take daily pills while others valued PrEP as a way to stay in control of their protection or remain HIV negative. At the interpersonal level, disclosure to partners and family could be difficult but support from friends and partners also encourage the use of PrEP. At the community level, stigma and myths about PrEP created a challenge yet public awareness campaigns and positive peer influence normalize it. At the institutional and organisational levels, barriers such as long queues, unfriendly staff and lack of confidentiality, along with inadequate information about PrEP were reported while access to PrEP information, youth friendly services as well as pharmacy-based models were seen as helpful. At the structural level, cost could act as a barrier but the availability of free PrEP at the point of care made it more accessible to many. These are described in more detail below.

### Barriers to accessing PrEP services

The results of the study identified barriers that align closely with the SEM, demonstrating how multilevel factors interconnect to influence young people’s access to, and use of, PrEP services (Table 2). At the individual level, difficulties adhering to daily pills and fear of needles reflect personal attitudes and behavioural limitations that constrain consistent use. Interpersonally, disclosure concern and fear of judgment reinforce stigma. Institutionally and organisational, long queues, negative staff attitudes, poor confidentiality and inadequate PrEP information discourage clinic engagement or visits. At the community level, the misconception that associates PrEP with HIV treatment further deter uptake. These factors collectively show how individual, interpersonal, institutional organisational as well as societal constraints converge to limit PrEP use.

**Table 2:**
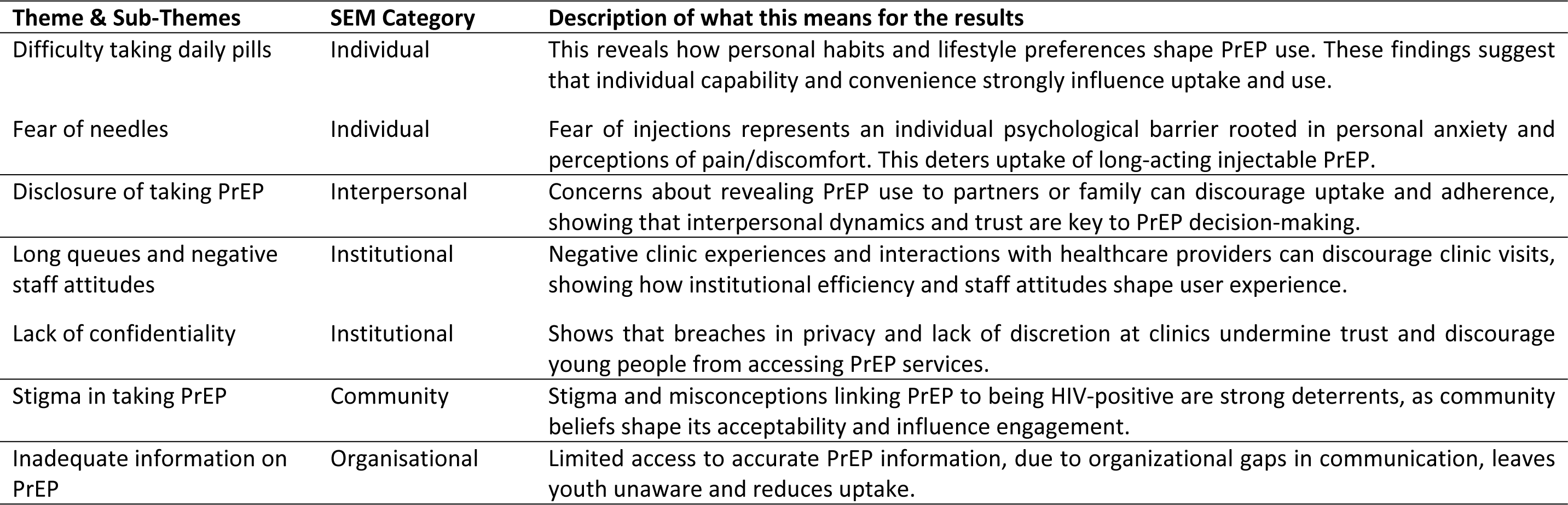
Barriers to accessing PrEP mapped to the Socio-Ecological Model (SEM)

Outlined below are the participants’ responses and perspectives as expressed during the study:

### Difficulty in taking a daily pill and Fear of needles (Individual level)

Adherence to a daily regimen was challenging, with some finding it difficult to take the pill daily. Young people expressed concerns about long-term adherence and the risk of forgetting the dosses as discouraging factors. As these participants noted:

> “Because the pills, I don’t like them, I wouldn’t be able to live taking them (pills) every day… yeah, so I prefer the injection…” (FGD04, Female, PrEP Naïve).

> “To drink it every day, because sometimes we make mistakes, like sometimes I would take 2 days, let’s say I forgot…I wouldn’t mind people knowing that I am taking PrEP, but what I mind for me is taking it every day… (FGD01, Male, PrEP Naïve).

Fear of needles emerged as an individual-level barrier to PrEP uptake among some youth. Participants explained that while injectable PrEP offers the advantage of long-term protection, many young people were hesitant due to a fear of needles. As one participant noted:

> “Some [youth] mostly don’t get along with the needles, yeah, some are scared…they are not going there because of the needles, the injections no… they prefer pills…”

Conversely, injectable PrEP was preferred by others who found daily pills burdensome and/or feared the stigma associated with being seen taking the medication regularly. One participant expressed a preference for injections to avoid daily medication:

> “Injection, I see it as the best option. It takes longer, and it is not something you would forget” (FGD05, Female PrEP Experienced).

> “Injection, it’s a once-off, a pill you forget. Injection once they do it, it is done (FGD05, Female PrEP Experienced).

However, participants also acknowledged the tension between fear and protection, recognizing that despite anxiety around fear of needles, prevention from HIV infection was highly valued.

> “…sometimes you are scared of the injection but you do want to prevent” (FGD01, Male, PrEP Naive).

### Disclosure of taking PrEP (Inter-personal level)

Participants expressed concern over disclosing taking PrEP, with some of the participants expressing anxiety about revealing their PrEP use to partners or family members.

> “You will drink it like, you wouldn’t want to show your person(partner) that look, I am eating these things [PrEP], she will ask what is that and it is for what?… now you have to explain that it is the pill …” (FGD01, Male, PrEP Naïve).

> “That is what I am running away from… like busy hiding. You know that we lose, and the sisters clean there at home, and then she finds it [pills] there…so eish!” (FGD01, Male, PrEP Naïve).

> “That thing ruins a relationship. How do you explain to your partner that you are taking that thing [PrEP]?”(FGD02, Male PrEP naïve).

### Long queues and negative staff attitudes at clinics (Institutional level)

Issues like long queues at clinics made regular visits time-consuming, and negative staff attitudes, especially judgmental or disrespectful behaviour, left many feeling unwelcome, further deterring them from accessing PrEP services. Together, these barriers highlight the difficulties young people face in navigating HIV prevention.

> “…the queues at the clinic are too much…like the line is too long and you can’t wait there knowing that you should go to work or do something you see” (FGD06, Male, PrEP Experienced).

> “The nurses, I think sometimes I don’t know if they are tired of people or what but eish their attitude most of the time…” (FGD03, Female, PrEP Naïve).

Pharmacies were highlighted for their convenience, shorter waiting times, and minimal interaction with others. This was especially valued by those seeking a more discreet and efficient way to access PrEP services. A participant shared:

> “As for me, I think pharmacy… because pharmacy could be simple, and then they open early, no queues, like again no more questions. So, at the clinic, sometimes there are people, so the queue, it is people who are sick, and all that, so as for me, I think pharmacy could be better…” (FGD03, Female, PrEP Naïve).

Others preferred pharmacies for their accessibility and the less judgmental environment, especially when compared to their previous negative experiences in clinics. One participant highlighted their preference for pharmacies, recalling an unpleasant experience at the clinic:

> “Yeah, I prefer pharmacy because the last time I went to the clinic, my father had HIV, so was there for the ARVs, and the nurse was shouting at him, saying it is not his date yet… and then we went back to the house…” (FGD04, Female PrEP Naïve).

### Lack of confidentiality (Institutional level)

A lack of perceived confidentiality was also a concern for study participants when visiting healthcare facilities for PrEP services. They expressed the need for confidentiality during consultations to feel secure and respected, especially given the sensitive nature of PrEP and its association with sexual health. Participants highlighted negative experiences at clinics, where a lack of discretion from healthcare providers made them uncomfortable and anxious. They felt that these breaches of confidentiality could lead to unintended disclosure of their reasons for visiting, which could be stigmatizing and embarrassing. One participant shared their experience:

> “Uhm, at the clinic, even when you are talking like she will shout you see, like the thing that you want to say is private, ‘speak loudly I cannot hear you,’ that time the other people will hear what you are here for…” (FGD03, Female PrEP Naïve).

### Stigma in taking PrEP (Community level)

Stigma emerged as the primary barrier to accessing PrEP services. Participants expressed hesitance in taking PrEP due to its association with antiretroviral drugs for HIV treatment. Taking daily pills was often perceived as a sign of living with HIV rather than an HIV preventive measure.

> “The other challenge could be stigma from the community, you see…or at home because now they will say I am like that [HIV positive] you see…yeah…even your person…” (FGD01, Male, PrEP Naive).

> “Us, the youth, we fear being looked by people……” (FGD06, Male, PrEP Experienced).

However, there was also a strong sentiment for making PrEP as easily accessible as condoms, which would help reduce stigma and increase availability. A male participant suggested:

> “I was thinking that they should be like condoms, anywhere, anyhow, so that you can remove the stigma about this thing.” (FGD02, Male, PrEP Naïve).

### Inadequate information about PrEP (Organisational level)

Inadequate information about PrEP was reported as one of the barriers to accessing PrEP. Many young people lack adequate information about PrEP, its benefits, and how it works. This knowledge gap prevents them from considering PrEP as a worthwhile option for HIV prevention.

> “I stopped it because they only gave me the PrEP, they didn’t explain because I went there, and I didn’t understand” (FGD05, Female, PrEP experienced).

> “The information no, I don’t have the whole information, I just know little bit” (FGD01, Female, PrEP Naive).

### Facilitators to accessing PrEP services

While participants described multiple barriers to accessing PrEP services, they also identified several factors that facilitated engagement with these services. These FGDs also identified several multilevel enablers that could strengthen PrEP uptake and sustained use (Table 3). Individually, a strong desire to remain HIV negative was a key motivator. Interpersonally and in the community, supportive relationships characterized by open communication and acceptance from family and friends promote health-seeking behavior and normalise PrEP use. Organisationally, youth-friendly information, respectful providers and convenient delivery models enhance access to PrEP. Awareness campaigns also reduce stigma while structural support, such as free PrEP provision, addresses the cost barriers.

**Table 3:**
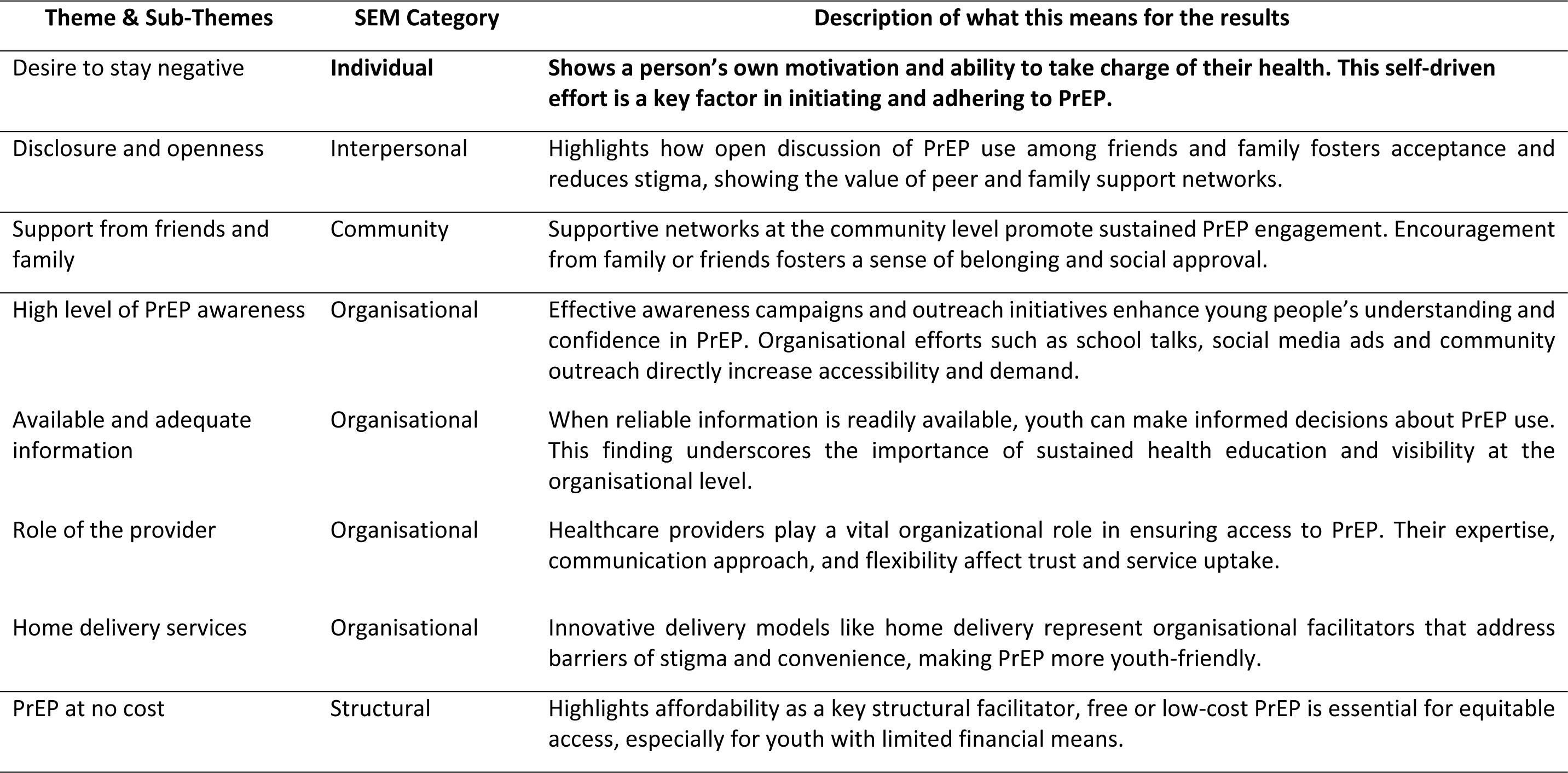
Facilitators to accessing PrEP mapped to the Socio-Ecological Model (SEM)

Outlined below are the participants’ responses and perspectives as expressed during the study:

### The desire to stay negative (Individual-level)

Participants expressed a strong personal motivation to remain HIV negative. The desire to “stay clean” shows a sense of responsibility for their own health and a proactive approach to HIV prevention.

> “Because I want to be clean always. That is my motivation to always take my medication. It is that I want to be clean.” (FGD05, Female, PrEP experienced).

### Disclosure (Inter-personal level)

Participants highlighted that openly disclosing the use of PrEP to friends and relatives raises a supportive environment that reduces stigma. This openness reduced the fear of being judged, instead it encourages acceptance.

> I think if I tell them at home that I am drinking this thing, they will give me food you see…” (FGD01, Male, PrEP Naïve).

### Support from friends and family (Community level)

Participants emphasized that support from friends and family also plays an important role, as social encouragement often reassures individuals and helps them feel more confident in their choice to use PrEP.

> “I would say support, like have that someone at home, like that support. Maybe to say, if they can see that I am taking my pill, they should tell me that, “did you take that thing(pills) of yours” and then I also become, committed, because as a person you cannot live with a sharp mind that reminds you…” (FGD04, Female PrEP Naïve).

### High level of PrEP awareness (Organisational level)

A high level of PrEP awareness among young people was seen as a facilitator at the organisational level. When organisations like clinics, pharmacies or NGOs, provided information through campaigns or outreach, participants reported that this will enable informed decision-making, empowering them to seek out PrEP services.

> “I think that if they are to say PrEP is being introduced, like every time, everywhere, where we walk, church, schools, clinics even at the mall, like as you walk at the entrance, there is a small tent the way those sisters that do HIV testing normally” (FGD03, Female PrEP Naïve).

> “I think they should advertise. Like, something that you see. It could be TV or billboards. Like, I also did not know about it before I got it. I didn’t know about it. I slept with someone, and I was worried. And then I came here. Before they gave me PrEP. They should do it on Facebook, maybe it is like an ad [advert] or something…” (FGD05, Female PrEP experienced)

### Available and adequate information (Organisational level)

Participants mentioned that the availability of information about PrEP ensures that those interested can easily access resources. Together, these factors facilitate greater engagement with PrEP services among young people.

> “But if you will give me information on why, like I hear that it would help me somewhere, I will try it for a day” (FGD02, Male, PrEP Naïve)

Some young people also demonstrated adequate knowledge of PrEP, understanding how it works and its benefits, which enhances their likelihood of accessing it. Participants further explained that much of what they knew about PrEP came from taking part in surveys or research studies rather than from a formal health education. They expressed a desire for more accessible information.

> “The more you busy doing surveys like this, like taking out information like this, for me one I have learned right, so for me it will be easy now, I now know the PrEP, …information… just take out the information……” (FGD01, Male, PrEP I).

### The role of the provider (Organisational level)

The insights suggest the importance of offering a range of healthcare providers for PrEP services to accommodate the varying preferences and comfort levels of young people. The results of the study highlighted the role of the providers as being significant in helping the young people access the PrEP services.

> “Because at the clinic, if I have the side effects … I will be able to talk to the sister so she checks me, like what is it that I am doing which makes me to be like that…” (FGD04, Female, PrEP Naïve)

Some participants preferred doctors due to their perceived higher level of expertise and comprehensive knowledge of PrEP and its effects. As one participant noted:

> “Doctor with more knowledge… yeah, doctors have more knowledge than the nurse…” (FGD05, Female, PrEP Experienced).

Others preferred a more flexible approach, where they would choose the healthcare provider based on the specific situation. For example, one participant expressed a preference for pharmacists when collecting PrEP, valuing the speed and efficiency of this interaction:

> “Yeah, that is when maybe I will want to see someone who counsels, but if I am collecting them then I can see a pharmacist, and then I pay, I leave.” (FGD05, Female, PrEP Experienced).

Meanwhile, some other young people were open to receiving services from a doctor or a nurse available, as long as they felt confident in the provider’s ability to assist with their needs:

> “Uhm with me, I prefer to be served by the doctor or nurse, both they are alright… as long as it will be a doctor or nurse…” (FGD06, Male, PrEP Experienced).

### Home delivery service (Organisational level)

Some participants found the convenience and discretion of home delivery services appealing. This option was seen as a way to avoid the potential stigma of being seen collecting PrEP at a healthcare facility. A participant shared their view on home delivery:

*“Delivery won’t show where it comes from. It will be discreet. They would have also called me that we are coming…” (FGDO2, Male PrEP Naïve)*.

### PrEP at no cost (Structural level)

Participants had a strong preference for affordable or free options, highlighting the importance of cost as a factor in accessing PrEP. Many participants expressed a desire for free access to PrEP, similar to how condoms are distributed for free in various locations, including clinics and shops. They felt that providing PrEP at no cost would encourage more people to use it and improve accessibility. One participant emphasized this point:

> “I think it will help if it is for free, and you get it from the clinic, even at the shops like I said, like even condoms can be found at the shops for free…” (FGD01, Male, PrEP Naïve).

Our findings show that young people face PrEP access barriers across all SEM levels with the strongest barrier at institutional and community levels which include long queues, stigma and poor confidentiality. Facilitators such as the desire to stay HIV-negative, supportive relationships and youth-friendly services helped encounter these barriers.

## Discussion

This study builds on the existing literature around barriers and facilitators to PrEP uptake in young people in sub-Saharan Africa. To our knowledge, it is also the first study to examine the barriers and facilitators related to accessing PrEP for both PrEP-experienced and PrEP-naive individuals and expands the age range of young people to 35 years, consistent with the South African definition[24]. This differs from the United Nations and many prior studies, such as a recent systematic review which define youth as 24 years and younger[25, 26]. Even though we conducted separate FGDs on these groups (naïve and experienced), the patterns of barriers and facilitators reported were consistently similar.

The study highlights a complex interchange or relationship of individual, interpersonal, community, institutional, structural and organisational level factors that shape young adults’ access to, and use of, PrEP services. These findings support similar multi-level analyses of PrEP uptake among young people in Uganda, Zimbabwe and South Africa[10, 27, 28]. Although the study population was stratified by PrEP experience and PrEP naïve, the barriers and facilitators to access PrEP were consistent across all groups. This suggests these multi-level factors are fundamental to the youth PrEP access and use experience regardless of prior PrEP exposure and experience.

At the individual level, the main barriers included difficulty taking the pill daily and fear of needles. Our results tell us that the challenge of taking a daily pill isn’t just about being forgetful, for others, it’s that they simply don’t want medication that requires that much daily effort. These findings align with other studies that also identified fear of needles and challenges with daily pill adherence as significant barriers, with some participants indicating they were unwilling to take the pill every day or afraid of injectable options[10, 28–30]. However, some indicated preference for injections over daily pills. This emphasizes the need for a tailored PrEP delivery model that meets young people’s preferences, including options for both oral pills and long-acting injectable forms[31].

In terms of interpersonal level factors, disclosure remains a sensitive issue, particularly with partners and family members. However, when young people disclosed and received encouragement, reminders to take pills or practical help with accessing the services, this support made it easier for them to stay on PrEP. This was echoed in another study conducted in Malawi showing that the uptake is generally high when provided with intensive support for initiation and adherence[32]. At the community level, misinformation about PrEP includes being mistaken for ARVs, as well as a lack of public awareness creates confusion and mistrust. Stigma remains a persistent obstacle, often worsened by poor confidentiality and judgmental attitudes in healthcare settings and in the community. These findings are consistent with existing literature, which shows that stigma can prevent youth from using HIV prevention tools like PrEP. In these studies, they emphasized community stigma as a big concern that creates a barrier to accessing PrEP[29, 33–35]. Combating community-level stigma requires public campaigns that normalize PrEP as part of comprehensive sexual health which is an approach endorsed by UNAIDS[36].

At the organisational level, one of the key barriers identified was inadequate information about PrEP. Many young people reported that they did not fully understand what PrEP is, how it works or its benefits. This lack of accessible information prevented them from making informed decisions or choices about whether to initiate (for the PrEP-naïve) or continue (for the PrEP-experienced) treatment. Inadequate information about PrEP was echoed in other studies, mentioning that healthcare provider interactions were pivotal in promoting PrEP uptake through the dissemination of accurate information and ongoing support[10, 28, 37]. This barrier was placed under the organisational level because it reflects how health facilities deliver information about PrEP. When these organisations fail to provide clear and accessible guidance, young people are left without the knowledge needed to start or remain on PrEP. This was mentioned by one PrEP-experienced participant who stopped taking PrEP because the healthcare worker did not give him information. In another study conducted in Kenya, their findings support the need for accurate and adequate information about PrEP to be provided to young people early on indecision making about PrEP use. This is likely to improve uptake of and adherence to PrEP[28, 38].

At the same time, the study shows that adequate information serves as a facilitator when organisations provide awareness campaigns, young people will feel empowered to seek PrEP services. In another study it was mentioned that providing information serves as a facilitator and can enhance engagement with PrEP among young people[39]. The role of the provider also came out as a facilitator with young people expressing the value to choose who and where they can receive services, whether doctors, nurses, pharmacies, clinics or home delivery options. This facilitator suggests youth-friendly service delivery models can enhance engagement with PrEP among young people.

On the institutional level, barriers associated with healthcare services were consistently reported, which include long queues, negative staff attitude and lack of confidentiality. Participants reported that healthcare workers as being judgmental towards young people. These experiences emphasize the need for youth-friendly services that are confidential and welcoming[40]. Alternative models of care like pharmacies and home deliveries were viewed as preferred and this could be leveraged to increase access of PrEP to young adults[41].

Finally, our findings show that many factors influencing PrEP use worked in ‘two-ways’, what acted as a barrier in one situation became a facilitator when conditions improved. For example, limited knowledge and poor privacy discouraged young people from initiating PrEP, but when they received clear information or experienced confidential, efficient services, these same areas motivated uptake. Recognizing these ‘two sides of the same coin’ suggest that improving PrEP uptake might not require new strategies, but rather strengthening the positive aspect of the existing services to directly counter the barriers youth face.

The findings of our study need to be considered in light of some limitations. We had a limited sample of PrEP experienced participants and while they brought rich, insightful contributions it is possible that they did not fully capture the experiences of this group more broadly. The findings of this study may also not be generalized to the broader population of young people across South Africa since the FGDs were conducted with a limited geographic area and may not fully represent the diversity of the larger target group in terms of socioeconomic status, education level, or experiences. Despite these limitations, this study provides unique insights into this important population residing within urban South Africa.

## Conclusion

This study highlights the multi-level factors influencing young adults’ access to PrEP, demonstrating how individual, interpersonal, community, organisational and structural factors interact to shape uptake and adherence. Young people face layered and intersecting barriers to accessing and using PrEP, shaped by individual concerns, social dynamics, health system constraints, and broader socio-ecological factors. While the desire to stay HIV-negative motivates many, stigma, limited knowledge, and unfriendly health service environments often undermine access. Our findings highlight the importance of offering tailored interventions that address whether it is structural or personal determinants of PrEP use, including comprehensive education campaigns, youth-friendly PrEP services and different PrEP modalities.

## Acknowledgments

We are particularly grateful to all the study B-Hub participants for the time and information they shared with us. The authors would like to thank the Indlela team for providing us with a list of potential participants. The authors declare no conflicts of interest.

## List of abbreviations

B-Hub: Behavioural Hub
FGD: Focus Group Discussions
GHSS: Global Health Sector Strategy
GP: Gauteng Province
HE²RO: Health Economics and Epidemiology Research Office
HIV: Human Immunodeficiency Virus
PLWH: People Living With HIV
PrEP: Pre-exposure Prophylaxis
SA: South Africa
SRH: Sexual Reproductive Health
SAA: Sub-Saharan Africa
UN-SDGs-3: United Nations Sustainable Development Goal 3
WHO: World Health Organization
SEM: Social-Ecological Models

## Consent for publication

Not applicable

## Availability of data and materials

The datasets used and/or analysed during the current study are available from the corresponding author upon reasonable request.

## Competing interests

The authors declare no competing interests.

## Funding statement

This research was funded by the Gates Foundation (INV-039268). LL was partially supported by the National Institute of Mental Health of the National Institutes of Health under grant number K01MH119923. The contents herein are the responsibility of the authors and do not necessarily reflect the views of the Gates Foundation or the National Institutes of Health.

## Authors’ contributions

CM and CH led the conception of the manuscript. CM led the design and drafted the manuscript, as well as conducted data analysis and interpretation of results. SD led the literature review. SD and MM. SD, MM, NM, RM contributed to the acquisition of the data, analysis and interpretation of the data, and critical revision of the manuscript. SB and CC-M contributed by helping with access to the sites and sample extraction. CH, LL, AM and JM contributed to overseeing the study and the critical revision of the manuscript. All authors reviewed and approved the final version of the manuscript.

## Notes

### Competing Interest Statement

The authors have declared no competing interest.

### Author Declarations

The study was conducted in compliance with regulatory requirements and ethical guidelines for research with human subjects in South Africa and institutional policies (ICH Good Clinical Practice). Ethical oversight was provided by the Human Research Ethics Committee (Medical) at the University of the Witwatersrand (211122), South African National Clinical Trials Registry, registration number: DOH-27-032022-5627 and the Gauteng Department of Health (GP_202201_028)

